# Temporal trends of SARS-CoV-2 seroprevalence in transfusion blood donors during the first wave of the COVID-19 epidemic in Kenya

**DOI:** 10.1101/2021.02.09.21251404

**Authors:** Ifedayo M.O. Adetifa, Sophie Uyoga, John N. Gitonga, Daisy Mugo, Mark Otiende, James Nyagwange, Henry K. Karanja, James Tuju, Perpetual Wanjiku, Rashid Aman, Mercy Mwangangi, Patrick Amoth, Kadondi Kasera, Wangari Ng’ang’a, Charles Rombo, Christine Yegon, Khamisi Kithi, Elizabeth Odhiambo, Thomas Rotich, Irene Orgut, Sammy Kihara, Christian Bottomley, Eunice W. Kagucia, Katherine E. Gallagher, Anthony Etyang, Shirine Voller, Teresa Lambe, Daniel Wright, Edwine Barasa, Benjamin Tsofa, Philip Bejon, Lynette I. Ochola-Oyier, Ambrose Agweyu, J. Anthony G. Scott, George M. Warimwe

## Abstract

Observed SARS-CoV-2 infections and deaths are low in tropical Africa raising questions about the extent of transmission. We measured SARS-CoV-2 IgG by ELISA in 9,922 blood donors across Kenya and adjusted for sampling bias and test performance. By 1st September 2020, 577 COVID-19 deaths were observed nationwide and seroprevalence was 9.1% (95%CI 7.6-10.8%). Seroprevalence in Nairobi was 22.7% (18.0-27.7%). Although most people remained susceptible, SARS-CoV-2 had spread widely in Kenya with apparently low associated mortality.

## Background

Across tropical Africa, numbers of cases and deaths attributable to COVID-19 have been substantially lower than those in Europe and the Americas. This could imply reduced transmission, reduced clinical severity or epidemiological under-ascertainment. The first COVID-19 case in Kenya was identified on 12^th^ March 2020. Subsequently, there have been two discrete waves of PCR-detected cases separated by a brief nadir in September 2020. At the end of 2020, the governement had recorded 96,458 cases and 1,670 deaths attributable to SARS-CoV-2. When COVID-19 related deaths reached 71, the national anti-SARS-CoV-2 antibody prevalence, estimated in blood donors, was 4.3% (95% confidence interval (CI) 2.9–5.8%) ^1^. Transmission was obviously more widespread than would have been anticipated by reported cases and deaths. In this further study, we examine the dynamics of SARS-CoV-2 seroprevalence among Kenyan blood donors throughout the course of the first epidemic wave.

## Methods

### Human samples

The Kenya National Blood Transfusion Service (KNBTS) coordinates and screens blood transfusion donor units at 6 regional centres at Eldoret, Embu, Kisumu, Mombasa, Nairobi and Nakuru, though the units are collected across the whole country and each Regional Centre serves between 5-10 of Kenya’s 47 Counties. KNBTS guidelines define eligible blood donors as individuals aged 16-65 years, weighing ≥50kg, with haemoglobin of 12·5g/dl, a normal blood pressure (systolic 120–129 mmHg and diastolic BP of 80–89 mmHg), a pulse rate of 60-100 beats per minute and without any history of illness in the past 6 months^2^. KNBTS generally relies on voluntary non-remunerated blood donors (VNRD) recruited at public blood drives typically located in high schools, colleges and universities. Since September 2019, because of reduced funding, KNBTS has depended increasingly on family replacement donors (FRD) who provide units of blood in compensation for those received by sick relatives. We obtained anonymized residual samples from consecutive donor units submitted to the 6 regional centres for transfusion compatibility-testing and infection screening, as previously described^1^.

### Laboratory analyses

We tested samples for anti-SARS-CoV-2 IgG antibodies using a previously described a previously described ELISA for whole length spike antigen^3^ at the KEMRI-Wellcome Trust Research Programme in Kilifi, Kenya. Assay sensitivity, estimated in sera from 174 PCR positive Kenyan adults and a panel of sera from the UK National Institute of Biological Standards and Control (NIBSC) was 92.7% (95% CI 87.9-96.1%); specificity, estimated in 910 serum samples from Kilifi drawn in 2018 was 99.0% (95% CI 98.1-99.5%)^1^. In a WHO-sponsored multi-laboratory study of SARS-CoV-2 antibody assays, results from Kilifi were consistent with the majority of the test laboratories^4^.

### Statistical analysis

We estimated crude prevalence based on the proportion of samples with OD ratio>2. We also used Bayesian Multi-level Regression with Post-stratification (MRP)^5^ to account for differences in the age and sex distribution of blood donors and regional differences in the numbers of samples collected over time. Data on donor residence were specified at County level. For the purposes of analysis and presentation we collapsed the 47 counties into 8 regions based on the previous administrative provinces of Kenya; as data from two regions (Eastern and North Eastern) was relatively sparse we collapsed these to one stratum. The model was also used to adjust for sensitivity (93%) and specificity (99%) of the chosen cut-off value as previously developed^6^. Regional and national estimates were produced by combining model predictions with weights from the 2019 Kenyan census^7^. Two versions of the model were fitted. In the first (Model A), the model included age, sex and region as covariates and was fitted separately to data in three periods (30 Apr–19 Jun, 20 Jun–19 Aug, 20 Aug–30 Sept). In the second (Model B), the model also included a period effect and was fitted to the samples as a whole. A mathematical description of the models and Rstan code^8^ is provided in the appendix.

### Ethical approval

This study was approved by the Scientific and Ethics Review Unit (SERU) of the Kenya Medical Research Institute (Protocol SSC 3426). Blood donors gave individual written consent for the use of their samples for research.

## Results

From 30^th^ April to 30^th^ September 2020, 10,258 blood donor samples were processed at six Kenya National Blood Transfusion Service (KNBTS) regional blood transfusion centres, which serve a countrywide network of satellites and hospitals. We excluded duplicate samples, those from age-ineligible donors and those with missing data, leaving 9,922 samples (Supplementary figure 1).

The blood donor samples were broadly representative of the Kenyan adult population^7^ on region of residence and age, although adults aged 55-64 years were under-represented (2.0% v 7.3%, Supplementary table 1) and adults aged 25-34 years were over-represented (39.3% v 27.3%). Males were also over-represented (80.8%).

Of the 9,922 samples with complete data 3,098 had been reported previously^1^. In total, 928 were positive for anti-SARS-CoV-2 IgG; crude seroprevalence was 9.4% (95% CI, 8.8-9.9%) with little variation by age or sex (Table 1a). We used Bayesian Multi-level Regression with Post-stratification (MRP) to adjust for test sensitivity (93%) and specificity (99%)^6^, smooth trends over time, and account for the differences in age, sex and residence characteristics of the test sample and the Kenyan population^5^.

**Table 1a.** Crude, age/sex standardized and Bayesian-weighted test-adjusted SARS-CoV-2 anti-spike protein IgG seroprevalence across the whole study duration.

|  | Kenya<br>population | All<br>samples | Sero-<br>positive | Crude<br>seroprevalence |  | Bayesian weighted, test-adjusted seroprevalence* |  |
| --- | --- | --- | --- | --- | --- | --- | --- |
|  |  |  |  | % | 95% CI | % | 95% CI |
| Age |  |  |  |  |  |  |  |
| 15 - 24 years | 9,733,174 | 2,763 | 241 | 8.7 | 7.7 - 9.8 | 7.5 | 6.2 - 8.8 |
| 25 - 34 years | 7,424,967 | 3,902 | 379 | 9.7 | 8.8 - 10.7 | 8.5 | 7.2 - 9.8 |
| 35 - 44 years | 4,909,191 | 2,261 | 224 | 9.9 | 8.7 - 11.2 | 8.3 | 6.9 - 9.8 |
| 45 - 54 years | 3,094,771 | 794 | 66 | 8.3 | 6.5 - 10.5 | 7.3 | 5.5 - 8.9 |
| 55 - 64 years | 1,988,062 | 202 | 18 | 8.9 | 5.4 - 13.7 | 7.2 | 5.2 - 9.1 |
| Sex |  |  |  |  |  |  |  |
| Male | 13,388,243 | 8,019 | 762 | 9.5 | 8.9 - 10.2 | 8.4 | 7.2 - 9.5 |
| Female | 13,761,922 | 1,903 | 166 | 8.7 | 7.5 - 10.1 | 7.4 | 5.9 - 8.9 |
| Region |  |  |  |  |  |  |  |
| Central | 3,452,213 | 606 | 38 | 6.3 | 4.5 - 8.5 | 5.8 | 3.7 - 8.0 |
| Coast | 1,671,097 | 1,680 | 137 | 8.2 | 6.9 - 9.6 | 7.2 | 5.6 - 8.9 |
| Eastern / N. Eastern | 5,176,080 | 1,482 | 108 | 7.3 | 6.0 - 8.7 | 6.5 | 4.9 - 8.2 |
| Mombasa | 792,072 | 1,654 | 239 | 14.4 | 12.8 - 16.2 | 13.8 | 11.7 - 16 |
| Nairobi | 3,002,314 | 607 | 107 | 17.6 | 14.7 - 20.9 | 16.7 | 13.4 - 20.2 |
| Nyanza | 3,363,813 | 1,433 | 131 | 9.1 | 7.7 - 10.8 | 8.3 | 6.6 - 10.2 |
| Rift Valley | 7,035,581 | 2,138 | 145 | 6.8 | 5.8 - 7.9 | 5.9 | 4.5 - 7.4 |
| Western | 2,656,995 | 322 | 23 | 7.1 | 4.6 - 10.5 | 6.6 | 3.9 - 9.7 |
| National | 27,150,165 | 9,922 | 928 | 9.4 | 8.8 - 9.9 | 7.9 | 6.7 - 9.0 |

\* Bayesian Multi-level Regression with Post-stratification (MRP) accounts for differences in the age and sex distribution of blood donors and regional differences in the numbers of samples collected over time. The model also adjusts for sensitivity (93%) and specificity (99%) of the ELISA

There was marked variation in seroprevalence over time and place with a generally increasing trend over time. Figure 1 illustrates the crude prevalence and Bayesian model estimates in 10 consecutive periods of approximately 2 weeks each. In Nairobi, Mombasa and the Coastal Region outside Mombasa, there was a steep rise in seroprevalence across the study period. We divided the observations equally into three consecutive periods (Table 1b). In period 1 (30 April-19 June) the adjusted seroprevalence of SARS-CoV-2 was 5.2% (95% CI 3.7-6.7%); in period 2 (20 June-19 August) it had risen to 9.1% (95% CI 7.2-11.3%); and in period 3 (20 August-30 September) it was maintained at 9.1% (95% CI 7.6-10.8%).

**Figure 1.**
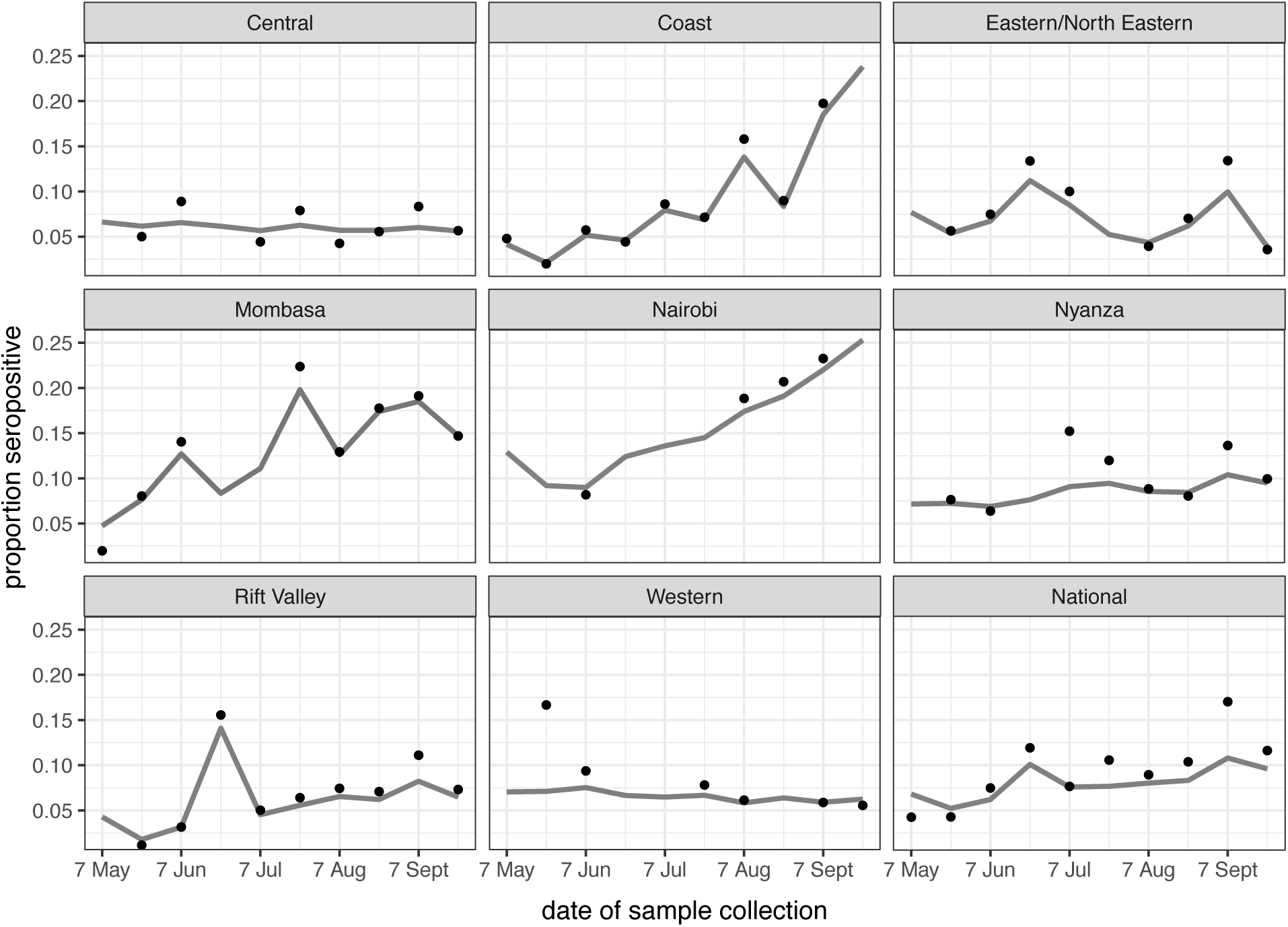
Seroprevalence positivity across the study period by region. The figure shows unadjusted estimates (black dots) and Bayesian model estimates (grey line) of seroprevalence in 8 regions of Kenya, and overall by date of sample collection in 10 periods of approximately 2 weeks each during 2020.

**Table 1b.** Bayesian-weighted test-adjusted SARS-CoV-2 anti-spike protein IgG seroprevalence across three study periods.

|  |  | Period 1 (30 Apr-19 Jun) |  | Period 2 (20 Jun-19 Aug) |  | Period 3 (20 Aug-30-Sep) |  |
| --- | --- | --- | --- | --- | --- | --- | --- |
|  |  | prevalence | 95%CI* | prevalence | 95%CI | prevalence | 95%CI |
| Age |  |  |  |  |  |  |  |
|  | 15 - 24 years | 5.2 | 3.4 - 7.1 | 8.3 | 5.9 - 10.8 | 8.7 | 6.9 - 10.6 |
|  | 25 - 34 years | 5.2 | 3.6 – 7.0 | 9.3 | 7.1 – 12.0 | 10.2 | 8.5 - 12.4 |
|  | 35 - 44 years | 6.1 | 4.1 - 8.6 | 10.5 | 7.7 - 13.8 | 8.7 | 6.8 - 10.7 |
|  | 45 - 54 years | 4.1 | 1.6 - 6.5 | 9.2 | 6.3 - 12.8 | 8.6 | 6.2 – 11.0 |
|  | 55 - 64 years | 4.1 | 1.1 - 6.9 | 8.7 | 4.6 - 12.9 | 8.8 | 6.3 - 12.9 |
| Sex |  |  |  |  |  |  |  |
|  | Male | 6.1 | 4.6 - 7.6 | 8.4 | 6.7 - 10.2 | 10.1 | 8.6 - 11.7 |
|  | Female | 4.3 | 2.3 - 6.5 | 9.7 | 6.9 - 13.1 | 8.2 | 6.1 - 10.5 |
| Region |  |  |  |  |  |  |  |
|  | Central | 5.3 | 2.5 – 9.0 | 6.9 | 3.9 - 10.4 | 5.9 | 3.0 - 9.5 |
|  | Coast | 3.3 | 1.9 - 4.8 | 11.3 | 7.4 – 16.0 | 14.1 | 10.6 - 18.1 |
|  | Eastern / N. Eastern | 5.4 | 3.2 - 8.2 | 9.6 | 6.7 - 12.9 | 5.2 | 3.4 - 7.3 |
|  | Mombasa | 7.4 | 5.0 - 10.1 | 17.2 | 12.5 – 23.0 | 15.9 | 13.1 - 19.1 |
|  | Nairobi | 6.7 | 3.9 - 10.5 | 10.1 | 3.7 - 20.1 | 22.7 | 18 - 27.7 |
|  | Nyanza | 5.3 | 3.2 - 7.7 | 11.3 | 8.0 - 15.2 | 8.5 | 6.1 - 11.3 |
|  | Rift Valley | 3.9 | 2.3 - 5.8 | 7.3 | 5.3 - 9.6 | 7.3 | 4.9 – 10.0 |
|  | Western | 6.3 | 2.9 - 11.6 | 7.7 | 3.9 - 12.2 | 6.0 | 2.4 - 11.1 |
|  | National | 5.2 | 3.7 - 6.7 | 9.1 | 7.2 - 11.3 | 9.1 | 7.6 - 10.8 |

\* 95% credible interval

## Discussion

The results illustrate a heterogeneous pattern of transmission across Kenya and suggest that the seroprevalence first began to rise in Mombasa in May and reached a maximum in July; in Nairobi it increased steadily from June onwards; in the Coastal area seroprevalence began to rise in July and turned up sharply in August and September. Unlike Nairobi and Mombasa this area is mostly rural. Other parts of the country showed less of a temporal trend. These field observations accord closely with epidemic modelling of SARS-CoV-2 across Kenya which integrated early PCR and serological data with mobility trends to describe the transmission pattern nationally^9^.

Although we used a highly specific and validated assay^1,4^, and adjusted for biases inherent in the ELISA test performance, we did not control for antibody waning. Given evidence at both individual^10^ and population^11^ level that anti-Spike antibodies may decline after an initial immune response, cross-sectional data are likely to underestimate cumulative incidence with increasing error as the epidemic wave declines. Some investigators have adjusted for this effect through modelling^12^ but as we do not have a clear description of the waning function for these antibodies in our setting, we have not made such an adjustment. Therefore, the seroprevalence estimates reported here are likely to underestimate cumulative incidence in Kenya.

The study also relies on convenience sampling of blood transfusion donors which is not representative of the adult population at large. We have adjusted for demographic and geographic disparities in our sample set but we are unable to evaluate whether the behaviour of blood donors makes them more or less at risk of infection by SARS-CoV-2. A random population sample would overcome this problem but such studies were difficult to undertake during movement restrictions. The selection bias in KNBTS samples is unlikely to change substantially over time and therefore this survey gives a valid estimation of trends, which inform the public health management of the epidemic.

The results are also consistent with other surveys in Kenya which have illustrated both high seroprevalence in focal populations and marked geographic variation. For example, seroprevalence was 50% among women attending ante-natal care (ANC) in August 2020 in Nairobi but 1.3%, 1.5% and 11.0% among women attending ANC in Kilifi (Coast) in September, October and November, respectively^13^. Seroprevalence among truck drivers at two sites (in Coast and Western) was 42% in October 2020^14^ and seroprevalence among health care workers in between August and November 2020 was 43%, 12% and 11%,in Nairobi, Busia (Western) and Kilifi (Coast) respectively^15^.

## Conclusion

By 1^st^ September 2020, the first epidemic wave of SARS-CoV-2 in Kenya had declined with a cumulative mortality of 577 COVID-19 deaths^16^. Our large national blood donor serosurvey illustrates that, at the same point, 1 in 10 donors had antibody evidence of infection with SARS-CoV-2; this rises to 1 in 5 in the two major cities in Kenya. The first epidemic wave rose and fell against a background of constant movement restrictions. The seroprevalence estimates suggest that population immunity alone was inadequate to explain this fall and the majority of the population remained susceptible. Nonetheless, they also show that the virus was widely transmitted during the first epidemic wave despite the fact that numbers of cases and deaths attributable to SARS-CoV-2 in Kenya were very low by comparison with similar settings in Europe and the Americas at similar seroprevalence^17,18^.

## Data Availability

The data shown in the manuscript are available upon request from the corresponding author. De-identified data has been published on the Havard dataverse server.

https://doi.org/10.7910/DVN/FQUNVD

## Acknowledgements

This project was funded by the Wellcome Trust (grant numbers 220991/Z/20/Z, 203077/Z/16/Z), the Bill and Melinda Gates Foundation (INV-017547) and by the Foreign Commonwealth and Development Office (FCDO) through the East Africa Research Fund (EARF/ITT/039). Sophie Uyoga is funded by DELTAS Africa Initiative [DEL-15-003], L. Isabella Ochola-Oyier is funded by a Wellcome Trust Intermediate Fellowship (107568/Z/15/Z), Ambrose Agweyu is funded by a DFID/MRC/NIHR/Wellcome Trust Joint Global Health Trials Award (MR/R006083/1), J. Anthony G. Scott is funded by a Wellcome Trust Senior Research Fellowship (214320) and the NIHR Health Protection Research Unit in Immunisation, Ifedayo Adetifa is funded by the United Kingdom’s Medical Research Council and Department For International Development through a African Research Leader Fellowship (MR/S005293/1) and by the NIHR-MPRU at UCL (grant 2268427 LSHTM). GMW is supported by a fellowship from the Oak Foundation. For the purpose of Open Access, the author has applied a CC-BY public copyright licence to any author accepted manuscript version arising from this submission.

This paper has been published with the permission of the Director, Kenya Medical Research Institute. We thank all of the blood donors for their contribution to the research.

## Competing interests

RA, MM, KK and PA are from the Ministry of Health, Government of Kenya. All other authors declare no competing interests.

## Data availability

The data shown in the manuscript are available upon request from the corresponding author. De-identified data has been published on the Havard dataverse server https://doi.org/10.7910/DVN/FQUNVD

## Notes

### Funding Statement

This project was funded by the Wellcome Trust (grant numbers 220991/Z/20/Z and 203077/Z/16/Z) the Bill and Melinda Gates Foundation (INV-017547) and by the Foreign Commonwealth and Development Office (FCDO) through the East Africa Research Fund (EARF/ITT/039). Sophie Uyoga is funded by DELTAS Africa Initiative [DEL-15-003]. L. Isabella Ochola-Oyier is funded by a Wellcome Trust Intermediate Fellowship (107568/Z/15/Z). Ambrose Agweyu is funded by a DFID/MRC/NIHR/Wellcome Trust Joint Global Health Trials Award (MR/R006083/1). J. Anthony G. Scott is funded by a Wellcome Trust Senior Research Fellowship (214320) and the NIHR Health Protection Research Unit in Immunisation. Ifedayo Adetifa is funded by the United Kingdom Medical Research Council and Department For International Development through a African Research Leader Fellowship (MR/S005293/1) and by the NIHR-MPRU at UCL (grant 2268427 LSHTM). GMW is supported by a fellowship from the Oak Foundation.

