## Supplementary for "Temporal trends of SARS-CoV-2 seroprevalence in transfusion blood donors during the first wave of the COVID-19 epidemic in Kenya"

### Supplementary Appendix

**Supplementary Figure 1. Participants flow diagram for SARS-CoV-2 seroprevalence study of blood donors in Kenya.**

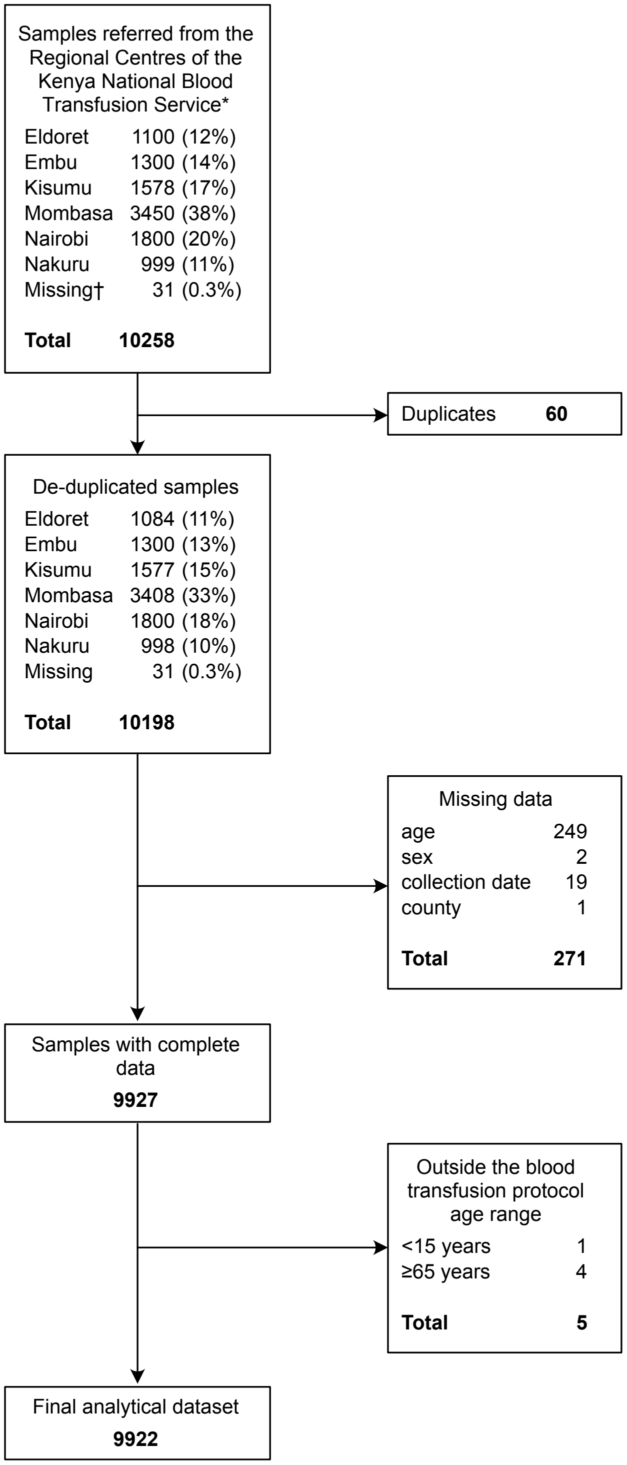


*3174 samples reported in an earlier publication are included in this analysis[1]
†All 31 records with missing information on Regional Centre were excluded further down the algorithm because they all lacked information on donor age.

**Supplementary Figure 2 Frequency of samples collected across the study period by region.**

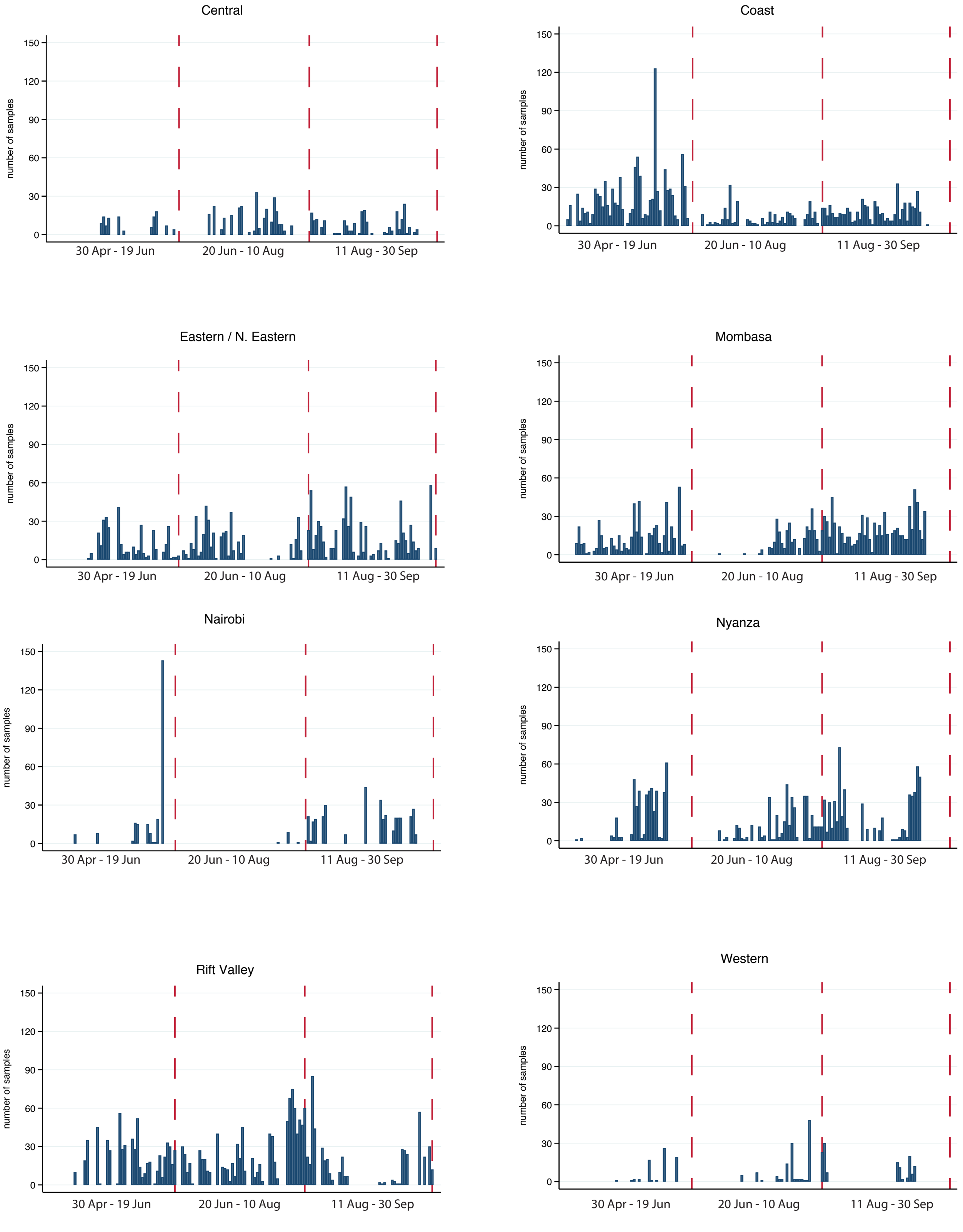

**Supplementary Table 1. General characteristics of the study population compared to the national population of Kenya.**

| Demographic variables | | Blood transfusion samples | | Kenya National Census 2019 | |
|---|---|---|---|---|---|
| | | N | % | N | % |
| Age | 15-24 years | 2,763 | 27.9 | 9,733,174 | 35.8 |
| | 25-34 years | 3,902 | 39.3 | 7,424,967 | 27.3 |
| | 35-44 years | 2,261 | 22.8 | 4,909,191 | 18.1 |
| | 45-54 years | 794 | 8.0 | 3,094,771 | 11.4 |
| | 55-64 years | 202 | 2.0 | 1,988,062 | 7.3 |
| Sex | Male | 8,019 | 80.8 | 13,388,243 | 49.3 |
| | Female | 1,903 | 19.2 | 13,761,922 | 50.7 |
| Region | Central | 606 | 6.1 | 3,452,213 | 12.7 |
| | Coast | 1,680 | 16.9 | 5,176,080 | 19.1 |
| | Eastern / N. Eastern | 1,482 | 14.9 | 792,072 | 2.9 |
| | Mombasa | 1,654 | 16.7 | 1,671,097 | 6.2 |
| | Nairobi | 607 | 6.1 | 3,002,314 | 11.1 |
| | Nyanza | 1,433 | 14.4 | 3,363,813 | 12.4 |
| | Rift Valley | 2,138 | 21.6 | 7,035,581 | 25.9 |
| | Western | 322 | 3.3 | 2,656,995 | 9.8 |
| | National | 9,922 | | 27,150,165 | |

N is the number of individuals in each stratum. % are column percentages

**Supplementary Table 2. A comparison of the general characteristics and SARS-CoV-2 seroprevalence in the blood donor populations in Kenya**

| | Family Replacement Donors | | | | Voluntary Non-Remunerated Donors | | | |
|---|---|---|---|---|---|---|---|---|
| | Number sampled | % of sample | Antibody positive | Seroprevalence (%) | Number sampled | % of sample | Antibody positive | Seroprevalence (%) |
| All donors | 9,632 | 100 | 899 | 9.3 | 290 | 100 | 29 | 10.0 |
| Male | 7,780 | 80.8 | 741 | 9.5 | 239 | 82.4 | 21 | 8.8 |
| Female | 1,852 | 19.2 | 158 | 8.5 | 51 | 17.6 | 8 | 15.7 |
| 15-24 years | 2,693 | 28.0 | 232 | 8.6 | 70 | 24.1 | 9 | 12.9 |
| 25-34 years | 3,793 | 39.4 | 368 | 9.7 | 109 | 37.6 | 11 | 10.1 |
| 35-44 years | 2,191 | 22.7 | 219 | 10.0 | 70 | 24.1 | 5 | 7.1 |
| 45-54 years | 765 | 7.9 | 64 | 8.4 | 29 | 10.0 | 2 | 6.9 |
| 55-64 years | 190 | 2.0 | 16 | 8.4 | 12 | 4.2 | 2 | 16.7 |

Voluntary Non-Remunerated Donors (VNRDs), who donate blood at community-based ‘blood drives’ comprised only 3% (290/9922) of our sample of donors during the pandemic; almost all donors in 2020 were Family Replacement Donors (FRDs) who provide a unit of blood in compensation for a transfusion received by a sick relative. Crude seroprevalence did not differ between the two groups.

**Supplementary Table 3. Bayesian weighted (unadjusted) and Bayesian weighted, test-performance adjusted seroprevalence estimates for the whole study period (30 April-30 Sept 2020), by sex, age and region.**

| | | Unadjusted seroprevalence | | Test-adjusted seroprevalence | |
|---|---|---|---|---|---|
| | | % | (95% CI) | % | (95% CI) |
| Sex | | | | | |
| | Male | 8.8 | 8.1-9.6 | 8.4 | 7.2-9.5 |
| | Female | 7.9 | 6.7-9.1 | 7.4 | 5.9-8.9 |
| Age | | | | | |
| | 15 - 24 years | 8.0 | 7.0-9.0 | 7.5 | 6.2-8.8 |
| | 25 - 34years | 8.9 | 8.0-9.9 | 8.5 | 7.2-9.8 |
| | 35 - 44 years | 8.7 | 7.8-9.9 | 8.3 | 6.9-9.8 |
| | 45 - 54 years | 7.9 | 6.4-9.1 | 7.3 | 5.5-8.9 |
| | 55 - 64 years | 7.8 | 6.1-9.4 | 7.2 | 5.2-9.1 |
| Region | | | | | |
| | Central | 6.4 | 4.7-8.4 | 5.8 | 3.7-8.0 |
| | Coast | 7.7 | 6.5-9.1 | 7.2 | 5.6-8.9 |
| | Eastern / N. Eastern | 7.1 | 5.9-8.5 | 6.5 | 4.9-8.2 |
| | Mombasa | 13.8 | 12.1-15.6 | 13.8 | 11.7-16.0 |
| | Nairobi | 16.4 | 13.6-19.6 | 16.7 | 13.4-20.2 |
| | Nyanza | 8.8 | 7.4-10.3 | 8.3 | 6.6-10.2 |
| | Rift Valley | 6.6 | 5.5-7.7 | 5.9 | 4.5-7.4 |
| | Western | 7.2 | 4.9-9.9 | 6.6 | 3.9-9.7 |
| | National | 8.4 | 7.6-9.1 | 7.9 | 6.7-9 |

**Supplementary Table 4. Study periods used in the analysis**

| Period | Start | Finish | Duration (days) | No. of samples | Median period date |
|---|---|---|---|---|---|
| 1 | 30 April 2020 | 19 June 2020 | 50 | 3362 | 02 June 2020 |
| 2 | 20 June 2020 | 19 August 2020 | 60 | 3837 | 27 July 2020 |
| 3 | 20 August 2020 | 30 September 2020 | 41 | 3723 | 01 September 2020 |

#### Statistical appendix:

Bayesian Multi-level Regression with Post-stratification (MRP) was used to account for differences in the age and sex distribution of blood donors and regional differences in the numbers of samples collected over time. This method involves fitting a hierarchical regression model and combining the resulting stratum-specific prevalence estimates with population weights to produce regional and national estimates. Because data on the sensitivity and specificity of the cut-off were also available, we were able to incorporate these parameters and use the model to estimate the "true" prevalence of seropositivity.

Two versions of the hierarchical regression model were fitted: one without a time period effect (Model A) and one with a period effect (Model B). Model A was fitted separately to samples in three periods (30 Apr – 19 Jun, 20 Jun – 19 Aug, 20 Aug – 30 Sept) and Model B was fitted to the combined data.

**Model A**

$$y_g = \text{Binomial}(n_g, p_g^*)$$

$$p_g^* = se \times p_g + (1 - sp) \times (1 - p_g)$$

$$\text{logit}(p_g) = \beta_{i[g]}^{sex} + \beta_{j[g]}^{region} + \beta_{k[g]}^{age}$$

$$\beta_i^{sex} \sim \text{Normal}(0, 10) \ \text{ for } i = 1, 2$$

$$\beta_j^{region} \sim \text{Normal}\left(0, \sigma_{region}\right) \ \text{ for } j = 1, .., 8$$

$$\beta_k^{age} \sim \text{Normal}(0, \sigma_{age}) \text{ for } k = 1, .., 5$$

$$\sigma_{region} \sim \text{Normal}^{+}(0, 0.5)$$

$$\sigma_{age} \sim \text{Normal}^{+}(0, 0.5)$$

$$\text{se} \sim \text{unif}(0, 1)$$

$$sp \sim \text{unif}(0, 1)$$

Region:

1 = central 2 = coast_mombasa 3 = coast_other 4 = eastern_neastern 5 = nairobi 6 = nyanza 7 = rift valley 8 = western

Sex:

1 = female 2 = male

Age group:

1 = 15-24yrs 2 = 25-34yrs 3 = 35-44yrs 4 = 45-54yrs 5 = 55-64yrs

Comments

-The true probability, $p_g$, of seropositivity in age-sex-region stratum $g$ is related to the observed probability, $p_g^*$ , through the equation $p_g^* = se \times p_g + (1 - sp) \times (1 - p_g)$.

-The effect of sex ($\beta_i^{sex}$) is modelled as a fixed effect and the effects of region ($\beta_{j[g]}^{region}$) and age ($\beta_{k[g]}^{age}$) are modelled as random effects. Modelling categorical variables as random effects can improve predictions for categories where only small amounts of data are available [6,19].

- The half-normal priors for $\sigma_{\text{age}}$ and $\sigma_{\text{region}}$ were chosen to be weakly informative [20]. To interpret these priors, note that if the baseline prevalence is 9% then SD = 0.5 on the logit scale corresponds to 95% of estimates being between 3.5% and 21.2%. Non-informative priors were used for all other parameters.

**Model B**

$$y_{gt} = \text{Binomial}(n_{gt}, p_{gt}^*)$$

$$p_{gt}^* = se \times p_{gt} + (1 - sp) \times (1 - p_{gt})$$

$$\text{logit}(p_{gt}) = \beta_{i[g]}^{sex} + \beta_{j[g]}^{region} + \beta_{k[g]}^{age} + \beta_{j[g],t}^{period}$$

$$\beta_{j,t}^{period} \sim \text{Normal}\big(\gamma_j(t-1), \sigma_{period,j}\big) \text{ for } t = 1,..,10$$

$$\gamma_j \sim \text{Normal}(0, \sigma_{slope})$$

$$\sigma_{period,j} \sim \text{Normal}^+(0, 0.5)$$

$$\sigma_{slope} \sim \text{Normal}^+(0, 0.05)$$

Comments

-The 10 periods correspond approximately to 2-week intervals of time.

-It is assumed that the period effects, $\beta_{j[g],t}^{period}$, follow a region-specific linear trend ($\gamma_j$). However, the model allows for some departure from a strict linear trend because the period effects are only constrained to follow a linear trend *on average*.

-The prior for $\sigma_{slope}$ is equivalent to that of the other SD parameters when the slopes are scaled to reflect change over the whole period.

**Model A**

```
data {
  int N_se; // denominator sensitivity
  int N_sp; // denominator specificity
  int x; // numerator sensitivity
  int z; // numerator specificity
  int y[5, 2, 8]; // no. seropositives
  int n[5, 2, 8]; // no. samples
  real pw[5, 2, 8]; // proportion of population in each demographic subgroup
  real tot_pw_age[5]; // proportion of population in each age group
  real tot_pw_sex[2]; // proportion female and male
  real tot_pw_region[8]; //proportion of population in each region
}

parameters {
  real<lower=0,upper=1> se;
  real<lower=0,upper=1> sp;
  real bsex[2];
  real bage[5];
  real bregion[8];
  real<lower=0> sd_age;
  real<lower=0> sd_region;
}

transformed parameters {
  real<lower=0,upper=1> p[5, 2, 8];
  real<lower=0,upper=1> p_obs[5, 2, 8];

  for(a in 1:5){
    for(s in 1:2){
      for(r in 1:8){
          p[a, s, r] = inv_logit(bage[a] +
                                 bsex[s] +
                                 bregion[r]);

           p_obs[a, s, r] = se * p[a, s, r] +
                              (1 - sp) * (1 - p[a, s, r]);
        }
      }
    }
}

model {
   //priors
   se ~ beta(1, 1);
```

```
  sp ~ beta(1, 1);
  bsex ~ normal(0, 10);
  bage ~ normal(0, sd_age);
  bregion ~ normal(0, sd_region);
  sd_age ~ normal(0, 0.5);
  sd_region ~ normal(0, 0.5);

 //likelihood

  for(a in 1:5){
   for(s in 1:2){
     for(r in 1:8){
          y[a, s, r] ~ binomial(n[a, s, r], p_obs[a, s, r]);
        }
      }
    }
   x ~ binomial(N_se, se);
   z ~ binomial(N_sp, sp);
}

generated quantities {
real p_national = 0;
vector[5] p_age = rep_vector(0, 5);
vector[2] p_sex = rep_vector(0, 2);
vector[8] p_region = rep_vector(0, 8);

   for(a in 1:5){
     for(s in 1:2){
       for(r in 1:8){
          p_national += p[a, s, r] * pw[a, s, r];
        }
      }
    }

   for(r in 1:8){
     for(a in 1:5){
       for(s in 1:2){
          p_region[r] += p[a, s, r] * pw[a, s, r]/tot_pw_region[r];
        }
      }
    }

   for(a in 1:5){
     for(r in 1:8){
       for(s in 1:2){
          p_age[a] += p[a, s, r] * pw[a, s, r]/tot_pw_age[a];
```

```
      }
    }
  }

  for(s in 1:2){
    for(a in 1:5){
      for(r in 1:8){
          p_sex[s] += p[a, s, r] * pw[a, s, r]/tot_pw_sex[s];
        }
      }
    }

}
```

**Model B:**

```
data {
  int N_se; //denominator sensitivity
  int N_sp; //denominator specificity
  int x; //numerator sensitivity
  int z; //numerator specificity
  int y[5, 2, 8, 10]; //no. of seropositives
  int n[5, 2, 8, 10]; //no. of samples
  real pw[5, 2, 8]; //proportion of population in each demographic subgroup
  real tot_pw_region[8]; //proportion of population in each region
}

parameters {
  real<lower=0,upper=1> se;
  real<lower=0,upper=1> sp;
  real bsex[2];
  real bage[5];
  real bregion[8];
  real bperiod[8, 10];
  real<lower=0> sd_age;
  real<lower=0> sd_region;
  real<lower=0> sd_period[8];
  vector[8] slope;
}

transformed parameters {
  real<lower=0,upper=1> p[5, 2, 8, 10];
  real<lower=0,upper=1> p_obs[5, 2, 8, 10];

  for(a in 1:5){
    for(s in 1:2){
```

```
      for(r in 1:8){
       for(t in 1:10){
          p[a, s, r, t] = inv_logit(bage[a] +
                                    bsex[s] +
                                    bregion[r] +
                                    bperiod[r, t]);

           p_obs[a, s, r, t] = se * p[a, s, r, t] +
                               (1 - sp) * (1 - p[a, s, r, t]);
          }
        }
      }
    }
}

model {
   //priors
   se ~ beta(1, 1);
   sp ~ beta(1, 1);
   bsex ~ normal(0, 10);
   bage ~ normal(0, sd_age);
   bregion ~ normal(0, sd_region);
   for(r in 1:8){
    for(t in 1:10){
      bperiod[r, t] ~ normal(slope[r] * (t - 1), sd_period[r]);
      }
    }
   sd_age ~ normal(0, 0.5);
   sd_region ~ normal(0, 0.5);
   sd_period ~ normal(0, 0.5);
   slope ~ normal(0, 0.05);

  //likelihood
  for(a in 1:5){
   for(s in 1:2){
    for(r in 1:8){
     for(t in 1:10){
          y[a, s, r, t] ~ binomial(n[a, s, r, t], p_obs[a, s, r, t]);
          }
        }
      }
    }
   x ~ binomial(N_se, se);
   z ~ binomial(N_sp, sp);
}
```

```
generated quantities {
vector[8] exp_slope = exp(slope);
vector[10] p_national = rep_vector(0, 10);
matrix[8, 10] p_region = rep_matrix(0, 8, 10);

  for(t in 1:10){
    for(a in 1:5){
      for(s in 1:2){
        for(r in 1:8){
           p_national[t] += p[a, s, r, t] * pw[a, s, r];
           }
        }
      }
    }

  for(t in 1:10){
    for(r in 1:8){
      for(a in 1:5){
        for(s in 1:2){
           p_region[r, t] += p[a, s, r, t] * pw[a, s, r]/tot_pw_region[r];
           }
        }
      }
    }

}
```
